# Sex and Life Stage Differences in the Association Between Cognitive Engagement and Cognitive Change

**DOI:** 10.64898/2026.09.11.26362841

**Authors:** Sarah Goulding, Shima Raeesi, John A. E. Anderson, Michael D. Oliver, Lisa L. Barnes, Mayra L. Estrella, Mahsa Dadar, Cassandra Morrison

## Abstract

**INTRODUCTION:** Cognitively stimulating activity (CSA) is associated with cognitive health, but whether associations with cognitive change differ by life stage and sex remains unclear.

**METHODS:** Data were drawn from Rush cohorts. Participants were 2,614 adults aged 65 years and older (1,965 females; 649 males). CSA during childhood, middle adulthood, and late life was classified using stage-specific pooled medians. Linear mixed-effects models examined global cognition and five domains, adjusting for age, education, and cognitive diagnosis. Sex-engagement-time interactions were tested with false discovery rate correction.

**RESULTS:** Childhood engagement showed limited associations; only the working-memory sex-engagement-time interaction survived correction (β = -0.014, *q*_FDR_=.047). Middle adulthood showed the strongest sex-dependent associations, with significant interactions for global cognition, episodic memory, semantic memory, processing speed, and visuospatial ability (*q*_FDR_≤.017). Higher engagement was associated with more favorable global and visuospatial trajectories among females but more negative trajectories across several domains among males. In late life, only working memory showed a significant sex interaction (β = 0.017, *q*_FDR_=.018). High engagement in childhood and middle adulthood followed by low late-life engagement was associated with more favorable global cognitive trajectories among females but less favorable trajectories among males compared with both consistently low and consistently high engagement (sex-pattern-time *q*_FDR_<.001).

*DISCUSSION:* Associations between CSA and cognitive change varied by life stage and sex and were not uniformly favorable. The observational design limits causal interpretation.

## 1. Introduction

Aging is associated with changes in global cognition as well as specific cognitive domains, including memory, language, attention, processing speed, and executive functioning^1,2^. However, the rate and extent of cognitive decline vary substantially across individuals and are influenced by both modifiable and nonmodifiable factors^3,4^. Engagement in cognitively stimulating activities (CSA), such as reading, writing, playing games, and other mentally demanding activities, has been proposed as a potentially modifiable contributor to cognitive health^5–7^. CSA may contribute to cognitive reserve, broadly defined as the capacity to maintain cognitive function in the presence of age-related brain changes or neuropathology^8–10^. Consistent with this framework, greater engagement in cognitively stimulating activities has been associated with better cognitive function, slower cognitive decline, and lower risk or later onset of dementia^11–15^.

Prior research has provided important evidence linking cognitive activity across the life course to cognitive aging and dementia^6,16–21^. Greater cognitive activity has been associated with a later age at onset of Alzheimer’s disease (AD) dementia^13^, while early-life cognitively enriching experiences have been associated with lower risk of mild cognitive impairment^14^. Other work in these cohorts has demonstrated associations between modifiable psychosocial factors and delayed dementia onset and has examined cognitive resilience in relation to AD and other neuropathologies^12,13^. Thus, the association between cognitive activity and cognitive aging is well established in several cohorts^6,13,16,17,22–24^. However, less is known about whether these associations differ by the life stage at which cognitive engagement occurs and whether these patterns vary by sex^25–28^. This question may be particularly relevant because engagement in CSA changes across the life course and may differ between females and males^25–27^. Engagement in CSA generally declines with age^2^, whereas older females have been reported to participate more frequently in cognitively stimulating leisure activities than males^2,25^.

Despite potential sex differences in both cognitive engagement and cognitive aging^25–28^, sex has not been consistently incorporated into cognitive reserve research^25–27^, highlighting an important gap in understanding whether associations between reserve-related factors and cognition differ by sex. Building on prior work^6,13,14,16,17^, the present study examined whether associations between CSA and longitudinal cognitive change differed by life stage and sex. CSA during childhood, middle adulthood, and late life, as well as selected lifelong engagement patterns, were evaluated. The primary focus was longitudinal change in global cognition, with additional analyses of episodic memory, semantic memory, working memory, processing speed, and visuospatial ability.

## 2. Methods

### 2.1 Participants

This study utilized data from the RADC Research Resource Sharing Hub. Participants were drawn from three ongoing longitudinal aging and dementia cohort studies: the Rush Memory and Aging Project (MAP)^24^, which began recruitment in 1997; the Minority Aging Research Study (MARS),^29^ which began recruitment in 2004; and the Latino Core (LATC),^23^ established later to extend recruitment to older Latino adults. Across these cohorts, standardized cognitive assessments, neuropsychological evaluations, and diagnostic classification procedures were harmonized to facilitate cross-cohort comparisons. Sex was recorded as female or male in the source cohort data; information on gender identity was not available.

Eligible participants were aged 65 years or older at baseline, had at least two cognitive assessment visits, had available baseline cognitive engagement data, and had a baseline clinical diagnosis. Participants with normal cognition, mild cognitive impairment, or dementia at baseline were retained in the primary analysis. After applying these criteria, the final analytic sample included 2,614 participants contributing 19,402 longitudinal observations. Cohort-specific characteristics and detailed inclusion/exclusion information are provided in Supplementary eTable 1. The parent cohort studies were approved by the Institutional Review Board of Rush University Medical Center, and all participants provided written informed consent. The studies were conducted in accordance with the Declaration of Helsinki.

### 2.2 Measures

#### 2.2.1 Cognitively Stimulating Activity

Childhood CSA was assessed using an 11-item composite measure reflecting the frequency of participation in cognitively stimulating activities at ages 6 and 12^18–21^. Participants reported engagement in 3 activities at age 6 and 8 activities at age 12, with each item rated on a 5-point scale. To distinguish cognitive engagement across discrete life stages, the childhood measure was restricted to activities reported at ages 6 and 12; activities at approximately age 18, representing young adulthood, were not included. Middle-adulthood CSA was evaluated using a 9-item self-report measure^30^ reflecting engagement in cognitively stimulating activities at approximately age 40 or during the decade from ages 30 to 40. Items were rated on a 5-point scale. Late-life CSA was assessed using a 7-item composite measure reflecting the frequency of participation in cognitively stimulating activities during the previous year, with each item rated on a 5-point scale.^31^ A detailed list of items for each life stage is provided in the Supplementary Materials.

#### 2.2.2 Cognitive Engagement Categorization

Participants’ engagement in CSA was categorized as high or low separately for childhood, middle adulthood, and late life. For each life stage, a single pooled median was calculated across the full eligible sample. The pooled median scores were 3.09 for childhood, 3.33 for middle adulthood, and 3.14 for late life. Participants with CSA scores at or above the median were classified as having high engagement, whereas those with scores below the median were classified as having low engagement. The same pooled median threshold was applied to females and males, and participants were subsequently grouped according to sex and engagement level. Engagement classifications were based on the baseline CSA assessment and were treated as fixed participant-level exposures in the longitudinal analyses.

Lifelong engagement patterns were defined from participants’ classifications across the three life stages. The prespecified patterns were high engagement across all three life stages (HHH; consistently high), low engagement across all three stages (LLL; consistently low), and high engagement in childhood and middle adulthood followed by low engagement in late life (HHL; high-high-low). These patterns were selected to contrast sustained high and low engagement with a pattern characterized by higher earlier-life and lower late-life engagement. The HHL group was compared separately with the LLL and HHH groups to examine whether lifelong engagement patterns were associated with subsequent cognitive trajectories.

#### 2.2.3 Cognitive Function

Cognitive testing was conducted at baseline and at annual follow-up assessments. Five cognitive domains were assessed using 19 cognitive tests: episodic memory, semantic memory, working memory, processing speed, and visuospatial ability. Individual test scores were standardized using the mean and standard deviation of the combined cohort at baseline and averaged within each domain to create domain-specific composite scores. A global cognition composite was derived by averaging standardized scores across the cognitive battery. Higher scores indicated better cognitive performance. A detailed description of the cognitive tests contributing to each domain is provided in the Supplementary Materials.

### 2.3 Statistical analysis

Data analyses were performed using *MATLAB R2024b*. Descriptive statistics were used to summarize participant characteristics overall and according to sex and cognitive engagement level. Continuous variables are presented as mean ± standard deviation, and categorical variables as counts and percentages, where applicable. Differences in demographic characteristics between high- and low-engagement groups were examined separately by sex using Welch two-sample *t* tests. Linear mixed-effects models were used to examine associations between CSA engagement during childhood, middle adulthood, and late life and longitudinal change in global cognition and five cognitive domains: episodic memory, semantic memory, working memory, processing speed, and visuospatial ability. For the present analyses, the resulting domain and global composite scores were z-scored across the pooled longitudinal analytic sample, whereas baseline age and education were z-scored across the participant-level baseline sample. Follow-up time was modeled continuously in years from baseline so that interaction coefficients represented differences in annual rates of cognitive change.

For each life stage, participants were classified into four sex-by-engagement groups (female-high, female-low, male-high, and male-low). Pairwise mixed-effects models compared longitudinal cognitive trajectories across these groups. The group-follow-up time interaction represented differences in annual cognitive change between groups. Models were adjusted for baseline age, years of education, and baseline cognitive diagnosis (normal cognition, mild cognitive impairment, or dementia). Participant-specific random intercepts were included to account for within-person correlation across repeated cognitive assessments. Analyses used available observations with complete data for all variables included in each model; missing values were not imputed. To formally evaluate whether the association between engagement and cognitive change differed by sex, separate models included a sex-engagement-follow-up time interaction for each life stage and cognitive outcome.

Lifelong engagement patterns were examined using global cognition as the outcome. The HHL group was compared separately with the LLL and HHH groups. Sex-specific pattern-time interactions were estimated, and sex-pattern-time interactions were used to formally test whether these associations differed between females and males. The primary analyses included eligible participants with normal cognition, mild cognitive impairment, or dementia at baseline. Because retrospective reports of earlier-life CSA may be less reliable among participants with dementia at baseline, all analyses were repeated after excluding participants with baseline dementia as a sensitivity analysis; these results are reported in the Supplementary Materials. As an additional sensitivity analysis, cognitive activity at approximately age 18 was combined with childhood activity to create an early-life engagement measure, consistent with prior RUSH studies^17,19,20^. The early-life score was calculated as the mean of the childhood and young-adulthood cognitive activity scores, and the primary childhood and lifelong analyses were repeated using this measure.

To account for multiple comparisons, Benjamini-Hochberg false discovery rate (FDR) correction^32^ was applied separately to prespecified families of tests. For the formal sex-engagement-time analyses, FDR correction was applied across the 18 tests representing three life stages and six cognitive outcomes. Pairwise group-time comparisons were corrected within each life stage across 36 tests, and lifelong analyses were corrected across six prespecified tests comprising sex-specific pattern-time effects and sex-pattern-time interactions for the two lifelong comparisons. FDR-adjusted *q* values <.05 were considered statistically significant.

## 3. Results

### 3.1 Demographics

The final analytic sample included 2,614 participants (1,965 females and 649 males). Females had a mean baseline age of 78.14 ± 7.31 years and a mean education of 14.42 ± 3.50 years, whereas males had a mean baseline age of 78.66 ± 7.04 years and a mean education of 15.43 ± 3.90 years. Mean follow-up was 6.92 ± 4.45 years for females and 6.39 ± 4.31 years for males. At baseline, 1,467 females and 434 males had normal cognition, 454 females and 187 males had mild cognitive impairment, and 44 females and 28 males had dementia (Table 1).

**Table 1.** Participant Characteristics by Sex.

|  | **Female** | **Male** |
| --- | --- | --- |
| Number of participants | 1965 | 649 |
| Age at Baseline | 78.14 ± 7.31 | 78.66 ± 7.04 |
| Mean Follow-up time (years) | 6.92 ± 4.45 | 6.39 ± 4.31 |
| Mean education (years) | 14.42 ± 3.50 | 15.43 ± 3.90 |
| Classification |  |  |
| NC | 1467 | 434 |
| MCI | 454 | 187 |
| Dementia | 44 | 28 |
| Baseline Global Cognition Composite Score | 0.017 ± 0.583 | -0.070 ± 0.624 |
| Mean Childhood Cognitive Engagement | 3.04 ± 0.72 | 2.92 ± 0.72 |
| Mean Middle Adulthood Cognitive Engagement | 3.25 ± 0.66 | 3.26 ± 0.68 |
| Mean Late Life Cognitive Engagement | 3.10 ± 0.71 | 3.02 ± 0.67 |

**Note.** NC = normal cognition; MCI = mild cognitive impairment. Global cognition is a composite score derived from standardized performance across the cognitive test battery, with higher scores indicating better cognitive performance. Values for number of participants and diagnostic classification are presented as n; all other values are mean ± SD.

Among the lifelong engagement patterns of primary interest, 549 females and 165 males were classified as HHH, 437 females and 170 males as LLL, and 167 females and 46 males as HHL. Mean age did not differ significantly between high- and low-engagement groups in childhood for either sex or in middle adulthood for either sex. In late life, participants in the high-engagement group were older than those in the low-engagement group among both females (*t* = 5.10, *p*<.001) and males (*t* = 3.51, *p*<.001). Educational attainment was higher in the high-engagement group than in the low-engagement group at all life stages for both sexes (all *p*<.001). Full demographic characteristics by sex and engagement level are presented in Supplementary eTables 2-4. Demographic characteristics and patterns across engagement groups were broadly similar after excluding participants with dementia at baseline (Supplementary eTables 5-9).

### 3.2 Cognitive Change

Formal sex differences in the association between cognitive engagement and longitudinal cognitive change were evaluated using sex-engagement-time interactions (Table 2). Middle adulthood showed the most consistent evidence of sex-dependent associations, with significant interactions after FDR correction for five of the six cognitive outcomes, whereas childhood and late-life interactions survived FDR correction only for working memory. Pairwise group-time comparisons were subsequently examined to characterize differences in cognitive trajectories between sex and engagement groups at each life stage (Tables 3-5). Adjusted global cognitive trajectories by sex and engagement during childhood, middle adulthood, and late life are shown in Figure 1. Lifelong engagement analyses also demonstrated sex-dependent differences in global cognitive trajectories, with adjusted trajectories for the two prespecified lifelong comparisons shown in Figure 2.

**Table 2.** Formal Sex-engagement-time Interactions Across the Three Life Stages.

|  | Global cognition | Episodic memory | Semantic memory | Working memory | Processing speed | Visuospatial ability |
| --- | --- | --- | --- | --- | --- | --- |
| **Childhood** | $\beta = -0.011$,<br>$t = -2.02$,<br>$p = .044$ | $\beta = -0.003$,<br>$t = -0.54$,<br>$p = .591$ | $\beta = -0.013$,<br>$t = -2.24$,<br>$p = .025$ | **$\beta = -0.014$,**<br>**$t = -2.36$,**<br>**$p = .018$** | $\beta = -0.006$,<br>$t = -1.13$,<br>$p = .257$ | $\beta = -0.015$,<br>$t = -2.11$,<br>$p = .035$ |
| **Middle Adulthood** | **$\beta = -0.022$,**<br>**$t = -4.15$,**<br>**$p < .001$** | **$\beta = -0.017$,**<br>**$t = -2.83$,**<br>**$p = .005$** | **$\beta = -0.021$,**<br>**$t = -3.61$,**<br>**$p < .001$** | $\beta = -0.006$,<br>$t = -0.92$,<br>$p = .359$ | **$\beta = -0.024$,**<br>**$t = -4.60$,**<br>**$p < .001$** | **$\beta = -0.020$,**<br>**$t = -2.83$,**<br>**$p = .005$** |
| **Late Life** | $\beta = 0.009$,<br>$t = 1.67$,<br>$p = .094$ | $\beta = 0.007$,<br>$t = 1.24$,<br>$p = .216$ | $\beta = 0.011$,<br>$t = 1.91$,<br>$p = .056$ | **$\beta = 0.017$,**<br>**$t = 2.74$,**<br>**$p = .006$** | $\beta = 0.003$,<br>$t = 0.56$,<br>$p = .576$ | $\beta = -0.002$,<br>$t = -0.27$,<br>$p = .790$ |

**Note.** $\beta$ represents the sex-engagement-time interaction coefficient. Female = 0, male = 1; low engagement = 0, high engagement = 1. Negative $\beta$ values indicate that the high-versus-low difference in annual cognitive change was more negative in males than females. Bolded values indicate statistical significance after false discovery rate (FDR) correction.

**Figure 1.**
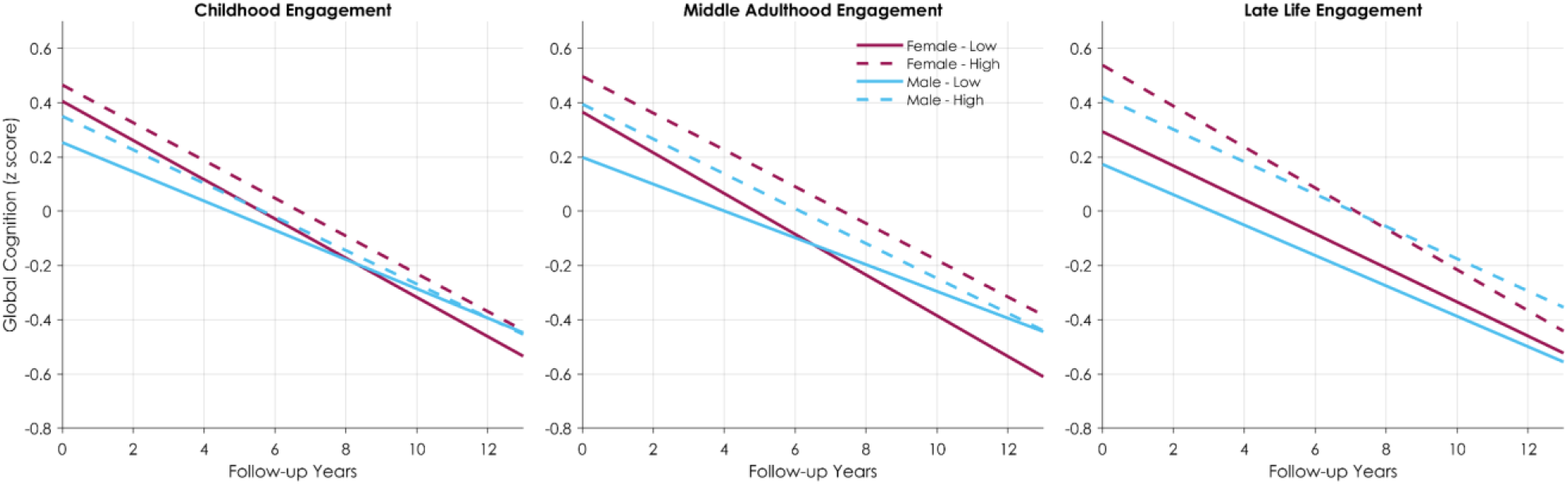
Cognitive Trajectories by Sex and Cognitive Engagement Across Life Stages. Predicted global cognitive trajectories are shown according to high versus low cognitively stimulating activity during childhood (left panel), middle adulthood (middle panel), and late life (right panel). Solid lines represent low engagement and dashed lines represent high engagement; female and male trajectories are shown separately. Predictions were derived from linear mixed-effects models adjusted for baseline age, education, and cognitive diagnosis, with normal cognition as the reference category. Follow-up was restricted to 12 years to reduce the influence of sparse observations at longer follow-up, and identical axis limits were used across panels.

### 3.3 Childhood

In childhood, the sex-engagement-time interaction was observed only for working memory after FDR correction (β = -0.014, *p* = .018, *q*_FDR_=.047), indicating that the association between childhood engagement and longitudinal working-memory change differed between females and males. No significant interactions were observed for global cognition, episodic memory, semantic memory, processing speed, or visuospatial ability after FDR correction (all *q*_FDR_>.05). Pairwise comparisons showed limited evidence of within-sex differences in cognitive trajectories according to childhood engagement. After FDR correction, no high-versus-low engagement comparison was significant among either females or males across the cognitive outcomes examined. Detailed pairwise estimates are presented in Table 3. The overall pattern was similar after excluding participants with dementia at baseline; however, the sex-engagement-time interaction for working memory no longer survived FDR correction (*q*_FDR_=.090; Supplementary eTable 10). Detailed pairwise sensitivity results are presented in Supplementary eTable 11. In the additional sensitivity analysis incorporating cognitive activity at approximately age 18, early-life associations remained limited and domain-specific; however, the significant sex-engagement-time interaction was observed for visuospatial ability rather than working memory (Supplementary eTable 12).

**Table 3.** Pairwise Group-Time Interactions for Childhood Cognitive Engagement.

|  | Global cognition | Episodic memory | Semantic memory | Working memory | Processing speed | Visuospatial ability |
| --- | --- | --- | --- | --- | --- | --- |
| **Female-high vs Female-low** | $\beta = 0.003$,<br>$t = 1.40$,<br>$p = .296$ | $\beta = -0.006$,<br>$t = -2.14$,<br>$p = .032$ | $\beta = 0.006$,<br>$t = 1.96$,<br>$p = .050$ | $\beta = 0.005$,<br>$t = 1.67$,<br>$p = .095$ | $\beta = 0.003$,<br>$t = 1.28$,<br>$p = .202$ | $\beta = 0.008$,<br>$t = 2.32$,<br>$p = .020$ |
| **Female-high vs Male-low** | $\beta = -0.016$,<br>$t = -4.21$,<br>$p < .001$ | $\beta = -0.017$,<br>$t = -4.05$,<br>$p < .001$ | $\beta = -0.016$,<br>$t = -4.04$,<br>$p < .001$ | $\beta = -0.006$,<br>$t = -1.37$,<br>$p = .170$ | $\beta = -0.002$,<br>$t = -0.63$,<br>$p = .529$ | $\beta = -0.003$,<br>$t = -0.68$,<br>$p = .494$ |
| **Female-high vs Male-high** | $\beta = -0.008$,<br>$t = -2.10$,<br>$p = .036$ | $\beta = -0.007$,<br>$t = -1.79$,<br>$p = .073$ | $\beta = -0.009$,<br>$t = -2.26$,<br>$p = .024$ | $\beta = 0.004$,<br>$t = 0.85$,<br>$p = .393$ | $\beta = 0.000$,<br>$t = 0.10$,<br>$p = .923$ | $\beta = 0.003$,<br>$t = 0.71$,<br>$p = .476$ |
| **Female-low vs Male-low** | $\beta = -0.018$,<br>$t = -4.93$,<br>$p < .001$ | $\beta = -0.011$,<br>$t = -2.60$,<br>$p = .009$ | $\beta = -0.022$,<br>$t = -5.34$,<br>$p < .001$ | $\beta = -0.011$,<br>$t = -2.46$,<br>$p = .014$ | $\beta = -0.006$,<br>$t = -1.50$,<br>$p = .133$ | $\beta = -0.011$,<br>$t = -2.24$,<br>$p = .025$ |
| **Female-low vs Male-high** | $\beta = -0.010$,<br>$t = -2.81$,<br>$p = .005$ | $\beta = -0.001$,<br>$t = -0.32$,<br>$p = .750$ | $\beta = -0.015$,<br>$t = -3.58$,<br>$p < .001$ | $\beta = -0.001$,<br>$t = -0.30$,<br>$p = .766$ | $\beta = -0.003$,<br>$t = -0.80$,<br>$p = .426$ | $\beta = -0.005$,<br>$t = -0.90$,<br>$p = .368$ |
| **Male-high vs Male-low** | $\beta = -0.008,$<br>$t = -1.90,$<br>$p = .058$ | $\beta = -0.009,$<br>$t = -1.95,$<br>$p = .051$ | $\beta = -0.007,$<br>$t = -1.63,$<br>$p = .103$ | $\beta = -0.009,$<br>$t = -1.83,$<br>$p = .067$ | $\beta = -0.003,$<br>$t = -0.61,$<br>$p = .542$ | $\beta = -0.007,$<br>$t = -1.13,$<br>$p = .258$ |

**Note.** $\beta$ represents the difference in annual cognitive change between the first-listed and second-listed groups. Positive $\beta$ values indicate a more positive (or less negative) annual cognitive trajectory in the first-listed group. Bolded values indicate statistical significance after false discovery rate (FDR) correction.

### 3.4 Middle Adulthood

Middle-adulthood engagement showed the strongest evidence of sex-dependent associations with cognitive change. Significant sex-engagement-time interactions after FDR correction were observed for global cognition (β = -0.022, *q*_FDR_<.001), episodic memory (β = - 0.017, *q*_FDR_=.017), semantic memory (β = -0.021, *q*_FDR_=.002), processing speed (β = -0.024, *q*_FDR_<.001), and visuospatial ability (β = -0.020, *q*_FDR_=.017). The interaction was not significant for working memory (*q*_FDR_=.431). In pairwise analyses, females with high middle-adulthood engagement showed more favorable trajectories than females with low engagement for global cognition and visuospatial ability after FDR correction. In contrast, among males, high engagement was associated with more negative trajectories in global cognition, episodic memory, semantic memory, and processing speed after FDR correction. Detailed pairwise comparisons are presented in Table 4. Results were essentially unchanged after excluding participants with dementia at baseline; sex-engagement-time interactions remained significant for global cognition, episodic memory, semantic memory, processing speed, and visuospatial ability, but not working memory (Supplementary eTable 10). Detailed pairwise sensitivity results are presented in Supplementary eTable 13.

**Table 4.** Pairwise Group-Time Interactions for Middle-Adulthood Cognitive Engagement.

|  | **Global cognition** | **Episodic memory** | **Semantic memory** | **Working memory** | **Processing speed** | **Visuospatial ability** |
| --- | --- | --- | --- | --- | --- | --- |
| **Female-high vs Female-low** | $\beta = \mathbf{0.007},$<br>$t = \mathbf{2.80},$<br>$p = \mathbf{.005}$ | $\beta = 0.003,$<br>$t = 1.18,$<br>$p = .237$ | $\beta = 0.006$<br>$t = 1.94,$<br>$p = .052$ | $\beta = 0.005,$<br>$t = 1.79,$<br>$p = .074$ | $\beta = 0.003,$<br>$t = 1.15,$<br>$p = .252$ | $\beta = \mathbf{0.009},$<br>$t = \mathbf{2.57},$<br>$p = \mathbf{.010}$ |
| **Female-high vs Male-low** | $\beta = \mathbf{-0.018},$<br>$t = \mathbf{-4.70},$<br>$p < \mathbf{.001}$ | $\beta = \mathbf{-0.016},$<br>$t = \mathbf{-3.64},$<br>$p < \mathbf{.001}$ | $\beta = \mathbf{-0.022},$<br>$t = \mathbf{-5.07},$<br>$p < \mathbf{.001}$ | $\beta = -0.001,$<br>$t = -0.21,$<br>$p = .836$ | $\beta = \mathbf{-0.014},$<br>$t = \mathbf{-3.55},$<br>$p < \mathbf{.001}$ | $\beta = -0.006,$<br>$t = -1.20,$<br>$p = .229$ |
| **Female-high vs Male-high** | $\beta = -0.004$,<br>$t = -1.04$,<br>$p = .299$ | $\beta = -0.002$,<br>$t = -0.63$,<br>$p = .530$ | $\beta = -0.006$,<br>$t = -1.67$,<br>$p = .095$ | $\beta = -0.001$,<br>$t = 0.15$,<br>$p = .884$ | **$\beta = 0.008$**<br>**$t = 2.25$**,<br>**$p = .024$** | $\beta = 0.005$,<br>$t = 1.10$,<br>$p = .272$ |
| **Female-low vs Male-low** | **$\beta = -0.026$**,<br>**$t = -6.35$**,<br>**$p < .001$** | **$\beta = -0.019$**,<br>**$t = -4.27$**,<br>**$p < .001$** | **$\beta = -0.027$**,<br>**$t = -6.21$**,<br>**$p < .001$** | $\beta = -0.006$,<br>$t = -1.36$,<br>$p = .175$ | **$\beta = -0.017$**,<br>**$t = -4.16$**,<br>**$p < .001$** | **$\beta = -0.015$**,<br>**$t = -2.74$**,<br>**$p = .006$** |
| **Female-low vs Male-high** | **$\beta = -0.011$**,<br>**$t = -3.00$**,<br>**$p = .003$** | $\beta = -0.006$,<br>$t = -1.43$,<br>$p = .154$ | **$\beta = -0.012$**,<br>**$t = -3.03$**,<br>**$p = .002$** | $\beta = -0.006$,<br>$t = -1.41$,<br>$p = .158$ | $\beta = 0.005$,<br>$t = 1.32$,<br>$p = .188$ | $\beta = -0.004$,<br>$t = -0.85$,<br>$p = .397$ |
| **Male-high vs Male-low** | **$\beta = -0.015$**,<br>**$t = -3.51$**,<br>**$p < .001$** | **$\beta = -0.013$**,<br>**$t = -2.74$**,<br>**$p = .006$** | **$\beta = -0.015$**,<br>**$t = -3.46$**,<br>**$p < .001$** | $\beta = 0.000$,<br>$t = -0.04$,<br>$p = .964$ | **$\beta = -0.021$**,<br>**$t = -4.87$**,<br>**$p < .001$** | $\beta = -0.011$,<br>$t = -1.81$,<br>$p = .071$ |

**Note.** $\beta$ represents the difference in annual cognitive change between the first-listed and second-listed groups. Positive $\beta$ values indicate a more positive (or less negative) annual cognitive trajectory in the first-listed group. Bolded values indicate statistical significance after false discovery rate (FDR) correction.

### 3.5 Late Life

In late life, a pattern similar to that observed in childhood emerged: the sex-engagement-time interaction remained significant after FDR correction only for working memory (β = 0.017, *p* = .006, *q*_FDR_=.018). No significant interactions were observed for global cognition, episodic memory, semantic memory, processing speed, or visuospatial ability after FDR correction. Among females, high late-life engagement was associated with more negative longitudinal trajectories than low engagement in global cognition, episodic memory, semantic memory, working memory, and processing speed after FDR correction, despite higher baseline global cognition in the high-engagement group. Among males, high- and low-engagement groups did not differ significantly in longitudinal cognitive change across any cognitive outcome. Adjusted global cognitive trajectories across the three life stages are shown in Figure 1, and detailed estimates are presented in Table 5. Results remained essentially unchanged after excluding participants with dementia at baseline, with working memory remaining the only domain showing a significant sex-engagement-time interaction after FDR correction (*q*_FDR_=.021; Supplementary eTable 10). Detailed pairwise sensitivity results are presented in Supplementary eTable 14.

**Table 5.** Pairwise Group-Time Interactions for Late-Life Cognitive Engagement.

|  | **Global cognition** | **Episodic memory** | **Semantic memory** | **Working memory** | **Processing speed** | **Visuospatial ability** |
| --- | --- | --- | --- | --- | --- | --- |
| **Female-high vs**<br>**Female-low** | $\beta = -0.013$,<br>$t = -4.71$,<br>$p < .001$ | $\beta = -0.008$,<br>$t = -2.84$,<br>$p = .004$ | $\beta = -0.013$,<br>$t = -4.30$,<br>$p < .001$ | $\beta = -0.014$,<br>$t = -4.53$,<br>$p < .001$ | $\beta = -0.009$,<br>$t = -3.26$,<br>$p = .001$ | $\beta = 0.001$,<br>$t = 0.42$,<br>$p = .677$ |
| **Female-high vs**<br>**Male-low** | $\beta = -0.019$,<br>$t = -4.61$,<br>$p < .001$ | $\beta = -0.013$,<br>$t = -2.77$,<br>$p = .006$ | $\beta = -0.020$,<br>$t = -4.44$,<br>$p < .001$ | $\beta = -0.006$,<br>$t = -1.20$,<br>$p = .231$ | $\beta = -0.009$,<br>$t = -2.25$,<br>$p = .024$ | $\beta = -0.003$,<br>$t = -0.56$,<br>$p = .577$ |
| **Female-high vs**<br>**Male-high** | $\beta = -0.016$,<br>$t = -4.64$,<br>$p < .001$ | $\beta = -0.012$,<br>$t = -3.14$,<br>$p = .002$ | $\beta = -0.019$,<br>$t = -5.18$,<br>$p < .001$ | $\beta = -0.009$,<br>$t = -2.35$,<br>$p = .019$ | $\beta = -0.003$,<br>$t = -0.95$,<br>$p = .343$ | $\beta = -0.002$,<br>$t = -0.58$,<br>$p = .559$ |
| **Female-low vs Male**<br>**low** | $\beta = -0.007$,<br>$t = -1.62$,<br>$p = .105$ | $\beta = -0.004$,<br>$t = -0.93$,<br>$p = .354$ | $\beta = -0.008$,<br>$t = -1.62$,<br>$p = .106$ | $\beta = 0.008$,<br>$t = 1.61$,<br>$p = .107$ | $\beta = 0.000$,<br>$t = -0.05$,<br>$p = .959$ | $\beta = -0.004$,<br>$t = -0.77$,<br>$p = .441$ |
| **Female-low vs Male**<br>**high** | $\beta = -0.003$,<br>$t = -0.93$,<br>$p = .352$ | $\beta = -0.003$,<br>$t = -0.86$,<br>$p = .387$ | $\beta = -0.006$,<br>$t = -1.64$,<br>$p = .101$ | $\beta = 0.005$,<br>$t = 1.21$,<br>$p = .264$ | $\beta = 0.006$,<br>$t = 1.51$,<br>$p = .130$ | $\beta = -0.004$,<br>$t = -0.85$,<br>$p = .393$ |
| **Male-high vs Male-**<br>**low** | $\beta = -0.003$,<br>$t = -0.79$,<br>$p = .431$ | $\beta = -0.001$,<br>$t = -0.21$,<br>$p = .833$ | $\beta = -0.001$,<br>$t = -0.27$,<br>$p = .789$ | $\beta = 0.004$,<br>$t = 0.68$,<br>$p = .494$ | $\beta = -0.006$,<br>$t = -1.28$,<br>$p = .199$ | $\beta = 0.000$,<br>$t = -0.06$,<br>$p = .949$ |

**Note.** $\beta$ represents the difference in annual cognitive change between the first-listed and second-listed groups. Positive $\beta$ values indicate a more positive (or less negative) annual cognitive trajectory in the first-listed group. Bolded values indicate statistical significance after false discovery rate (FDR) correction.

### 3.6 Lifelong Engagement Patterns

Lifelong engagement patterns showed pronounced sex differences in global cognitive trajectories. Compared with LLL, HHL was associated with a more favorable global cognitive trajectory among females (β = 0.038, *q*_FDR_<.001) but a less favorable trajectory among males (β = -0.049, *q*_FDR_<.001). The corresponding sex-pattern-time interaction was significant (β = -0.088, *q*_FDR_<.001). A similar sex-dependent pattern was observed for the HHL versus HHH comparison: HHL was associated with a more favorable trajectory among females (β = 0.034, *q*_FDR_<.001) but a less favorable trajectory among males (β = -0.038, *q*_FDR_=.001), with a significant sex-pattern-time interaction (β = -0.073, *q*_FDR_<.001). Adjusted global cognitive trajectories for both comparisons are shown in Figure 2. Lifelong engagement findings were also robust to exclusion of participants with dementia at baseline; the sex-specific pattern-time effects and sex-pattern-time interactions remained significant for both comparisons (Supplementary eTable 15). Lifelong findings were also directionally consistent when the age-18 cognitive activity measure was incorporated into the early-life classification, with significant sex-specific pattern-time effects and sex-pattern-time interactions for both comparisons (Supplementary eTable 16).

**Figure 2.**
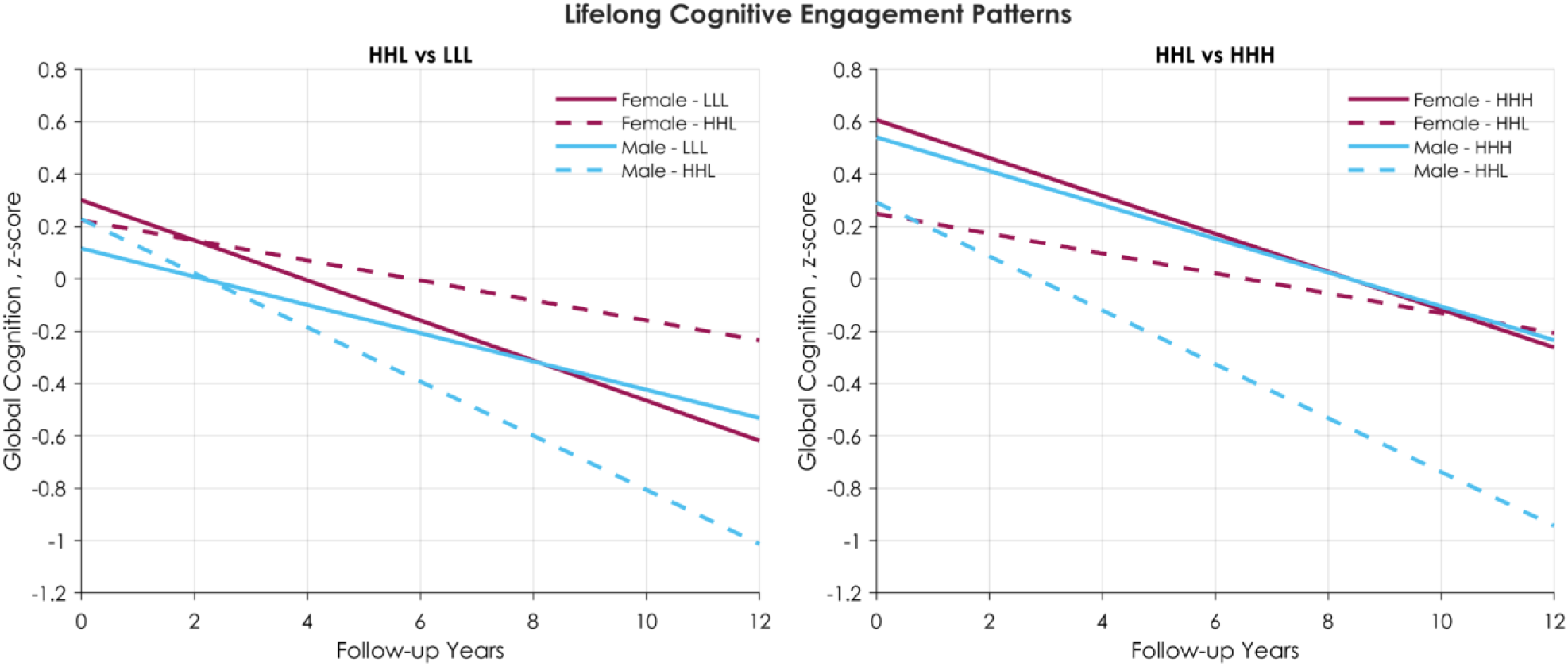
Global Cognitive Trajectories by Sex and Lifelong Cognitive Engagement Pattern. Predicted global cognitive trajectories are shown for participants with high engagement in childhood and middle adulthood followed by low engagement in late life (HHL; high-high-low), compared with those with low engagement across all three life stages (LLL; low-low-low; left panel) and high engagement across all three life stages (HHH; high-high-high; right panel). Solid lines represent the comparison groups (LLL or HHH), and dashed lines represent HHL; female and male trajectories are shown separately. Predictions were derived from linear mixed-effects models adjusted for baseline age, education, and cognitive diagnosis, with normal cognition as the reference category. Follow-up was restricted to 12 years to reduce the influence of sparse observations at longer follow-up, and identical y-axis limits were used across panels.

## 4. Discussion

This study examined whether associations between CSA and longitudinal cognitive change varied by life stage and sex. Middle adulthood showed the strongest sex-dependent associations, with higher engagement linked to more favorable trajectories among females but more negative trajectories among males. Childhood associations were limited, whereas late-life and lifelong engagement showed more complex patterns.

To our knowledge, few longitudinal studies^26,29,33^ have simultaneously examined whether associations between cognitive engagement and cognitive change vary by both life stage and sex across multiple cognitive domains. The present study extends previous work by considering life stage, sex, cognitive domain, and lifelong engagement patterns within the same longitudinal framework. Prior studies have demonstrated associations between cognitive activity across the life course and cognitive aging^6,13,14,16–19^, while other research has highlighted the limited incorporation of sex and gender into cognitive reserve research^25–27^. Earlier- and later-life cognitive activity have both been associated with slower cognitive decline^16–18,31,34,35^, whereas relatively few cognitive reserve studies have examined whether these associations differ by sex^26,29,33^. Thus, the present findings suggest that associations between CSA and subsequent cognitive trajectories may depend not only on the amount of engagement, but also on when engagement occurs and on sex. This is particularly relevant given growing evidence that sex- and gender-related factors may modify resilience mechanisms across aging and AD stages^26,29,33,36^.

The relatively weak findings for childhood engagement are consistent with some prior studies suggesting that early-life cognitive enrichment may be more strongly related to later-life cognitive level than to subsequent decline^37^. In one Rush cohort study, foreign-language and music instruction during childhood and adolescence was associated with higher late-life cognition and reduced risk of mild cognitive impairment, but not with subsequent cognitive decline^37^. This closely parallels the limited within-sex trajectory differences observed in the present study. However, other work from Rush cohorts reported that greater early-life cognitive activity was associated with slower late-life cognitive decline even after accounting for neuropathologic burden^17^. Differences across studies may reflect variation in exposure definitions, continuous versus categorical modeling, cognitive outcomes, and clinical composition^17,37^. Childhood CSA was also recalled many decades later, which may introduce measurement error and attenuate associations^17,38^. Given that the only significant childhood sex interaction did not remain after exclusion of participants with dementia, the childhood findings should be interpreted cautiously. The limited nature of these associations was also evident when cognitive activity at approximately age 18 was incorporated into an early-life measure, although the specific cognitive domain showing a sex-dependent association differed.

In contrast, middle adulthood emerged as the clearest period of sex-dependent association. Among females, higher engagement was associated with more favorable global cognitive and visuospatial trajectories. This pattern is consistent with evidence linking midlife cognitive activity to later cognitive health^7,11,12,14,31,39^. In a 44-year longitudinal study of women, greater cognitive activity in midlife was associated with lower risks of both dementia and AD^40^. Among males, however, higher engagement was associated with more negative trajectories in global cognition, episodic memory, semantic memory, and processing speed. This finding should not be interpreted as evidence that CSA is harmful, but may instead reflect heterogeneity in what similar activity-frequency scores represent for females and males^25,26,33^. Women and men may differ in both their activity profiles and the associations of specific activities with later cognitive outcomes^25,26,33,41^. For example, longitudinal studies suggest that associations between adulthood leisure activities and subsequent cognitive trajectories may vary by activity type and sex^25,28,41^. Gender-related differences in occupational demands, educational and socioeconomic opportunities, caregiving, stress exposure, and the qualitative complexity of activities may therefore contribute to the observed sex differences^42–44^. Prior cognitive reserve research similarly emphasizes that conventional reserve proxies are shaped by sex- and gender-related social experiences^26,33^.

The late-life findings were more unexpected. Previous studies have generally reported that greater cognitive activity in later life is associated with better cognitive outcomes^6,11,15,16,31^. Rush cohort studies have similarly linked greater late-life activity with better subsequent cognitive functioning and slower decline^17,19,31^. In the present study, however, females with high late-life engagement had higher baseline global cognition but subsequently showed steeper decline in global cognition, episodic memory, semantic memory, working memory, and processing speed. One possible explanation is compatible with a cognitive reserve pattern in which individuals maintain higher cognitive performance despite accumulating pathology but decline more rapidly once compensatory capacity is exceeded^8,9,45^. Previous longitudinal studies provide some support for this interpretation. Greater cognitive activity has been associated with a later onset of accelerated memory decline among individuals who subsequently developed dementia^14,30^, while slower decline before dementia onset but faster decline after dementia becomes clinically manifest has also been reported^30,34,46–50^. Similar reserve-related patterns have also been described for educational attainment^48,49,51,52^.

However, this interpretation cannot be confirmed from the present data and should be considered preliminary. The late-life association persisted after participants with baseline dementia were excluded, suggesting that it was not driven solely by diagnosed dementia at study entry. Nevertheless, participants with normal cognition or mild cognitive impairment may still have had substantial preclinical neuropathology^9,53–55^. Late-life activity is also particularly susceptible to reverse causation and health-related selection because engagement may change in response to emerging cognitive, functional, social, or health changes^17,31,56^. Importantly, although several female high-versus-low trajectory differences survived FDR correction, the sex-engagement-time interaction was significant only for working memory. Thus, the strongest evidence for a sex difference in late-life associations was specific to working memory rather than all cognitive domains.

The lifelong engagement findings further suggest that cognitive engagement cannot be understood simply as a cumulative exposure in which greater engagement at every stage necessarily predicts slower cognitive decline^8,9^. Among females, the HHL pattern was associated with more favorable global cognitive trajectories than both LLL and HHH, whereas among males the same pattern was associated with less favorable trajectories. Although previous research has generally supported beneficial associations of cognitive activity accumulated across multiple life stages^17,19,31,57^, the present findings suggest that the timing and pattern of engagement may matter differently by sex. Among females, the more favorable HHL versus LLL trajectory may be compatible with a lasting contribution of greater earlier-life engagement^17,19,37^. However, the more favorable HHL versus HHH trajectory should not be interpreted as evidence that lower late-life engagement is beneficial. It may instead reflect the higher-baseline/faster-decline pattern observed in the late-life analysis or differences in the circumstances underlying distinct life-course engagement patterns^5,8,9,45,56^. Retirement, caregiving demands, health changes, occupational histories, and social opportunities may influence lifelong engagement differently in females and males^25,26,33,42^.

Taken together, these findings may help explain heterogeneity in the cognitive-engagement literature. Some studies have found associations between greater cognitive activity and slower cognitive decline or lower dementia risk^6,12,14,16,17,31,39^, whereas others have reported stronger associations with baseline or attained cognitive level than with subsequent rate of change^28,37,58^. The present results suggest that some of this heterogeneity may reflect life stage, cognitive domain, and sex. These findings suggest that associations between CSA and cognitive aging may vary in direction and magnitude across life stages and sexes^9,26,33^. This interpretation is consistent with work emphasizing that cognitive resilience reflects interacting biological and social determinants whose effects may vary by sex, age, and disease stage^9,26,33,59–61^.

This study has several strengths, including the large longitudinal sample, repeated annual cognitive assessments, multiple cognitive domains, life-course CSA measures, formal sex-interaction testing, and sensitivity analyses excluding baseline dementia. Several limitations should also be considered. Childhood and middle-adulthood CSA were assessed retrospectively and may therefore be affected by recall error^17,38^, particularly given the long interval between the activities and their assessment. Self-reported activity measures are also susceptible to social desirability and may not capture cognitive complexity, intensity, or social context^25,26,62^. The use of median splits facilitates interpretation but reduces information and may obscure nonlinear associations^63^. Late-life engagement was classified at baseline and treated as a fixed exposure, although engagement may change substantially during follow-up^25,31^. Given the observational design, causal relationships cannot be established, and unmeasured socioeconomic, occupational, health, social, caregiving, and other life-course factors may have influenced the observed associations^3,4,26,33,42^. Finally, because measures of neuropathology or neuroimaging were not incorporated into these analyses, the observed patterns cannot be attributed directly to cognitive reserve mechanisms.

In conclusion, associations between cognitive engagement and cognitive aging varied by life stage and sex. Middle adulthood showed the strongest sex-dependent associations, childhood findings were comparatively weak, and late-life and lifelong analyses showed more complex sex-specific patterns. These findings suggest that the timing and pattern of cognitive engagement may contribute to heterogeneity in cognitive aging and underscore the importance of considering sex and gender-related life-course experiences.

## Acknowledgments

The authors thank the participants of the Rush Memory and Aging Project (MAP), the Minority Aging Research Study (MARS), and the Rush Alzheimer’s Disease Center (RADC) Latino Core, as well as the staff of the Rush Alzheimer’s Disease Center, for their invaluable contributions and commitment.

## Data Availability Statement

The data analyzed in this study are available through the Rush Alzheimer’s Disease Center [RADC Research Resource Sharing Hub](https://www.radc.rush.edu/). Access is subject to RADC data-sharing procedures and applicable data use requirements.

